# Pathways mediating the effect of education on pregnancy outcomes: A Mendelian randomization study

**DOI:** 10.1101/2023.07.07.23292364

**Authors:** Tormod Rogne, Dipender Gill, Zeyan Liew, Xiaoting Shi, Vilde Hatlevoll Stensrud, Tom Ivar Lund Nilsen, Stephen Burgess

## Abstract

**Objective:** To investigate the relationship between education and pregnancy outcomes, and the proportion of the effect of education mediated through modifiable cardiometabolic risk factors, using two-sample Mendelian randomization (MR) analyses.

**Methods and Analysis:** We extracted uncorrelated (*R*^2^ <0.01) single-nucleotide polymorphisms strongly associated (p-value <5e-8) with educational attainment, type 2 diabetes mellitus, body mass index, smoking, high-density lipoprotein cholesterol, and systolic blood pressure from the largest genome-wide association studies with available summary data. Genetic associations with ectopic pregnancy, hyperemesis gravidarum, gestational diabetes, preeclampsia, preterm birth, and offspring birth weight were extracted from the largest genome-wide association studies with available summary data. All subjects were of European ancestry. We conducted univariable MR analyses with the inverse-variance weighted method employed in the main analysis, and weighted median, weighted mode and MR Egger regression in the sensitivity analyses to account for potential pleiotropy. In mediation analyses, we compared the direct effect of educational attainment estimated in multivariable MR with the total effect estimated in the main univariable MR analysis.

**Results:** The analyses included more than 3 million subjects with data on educational attainment, 270,002 subjects with data on offspring birth weight, and between 2,092 and 15,419 cases with adverse pregnancy outcomes. Each standard deviation increase in genetically-predicted educational attainment (3.4 years) was associated with an increased birth weight (95% confidence interval) of 42 g (28 g to 56 g) and an odds ratio (95% confidence interval) of 0.53 (0.46 to 0.60) for ectopic pregnancy, 0.54 (0.44 to 0.66) for hyperemesis gravidarum, 0.73 (0.67 to 0.80) for gestational diabetes, 0.81 (0.71 to 0.93) for preeclampsia, and 0.72 (0.67 to 0.77) for preterm birth. The combined proportion of the effect (95% confidence interval) of genetically-predicted educational attainment that was mediated by the five cardiometabolic risk factors was 42% (14% to 59%) for ectopic pregnancy, -17% (-46% to 26%) for hyperemesis gravidarum, 48% (19% to 82%) for gestational diabetes, 78% (10% to 208%) for preeclampsia, 28% (0% to 51%) for preterm birth, and 9% (-26% to 24%) for birth weight. Sensitivity analyses accounting for pleiotropy were consistent with the main analyses.

**Conclusion:** Our findings support that intervening on type 2 diabetes mellitus, body mass index, smoking, high-density lipoprotein cholesterol, and systolic blood pressure would lead to reductions in several adverse pregnancy outcomes attributable to lower levels of education. Such public health interventions would serve to reduce health disparities attributable to social inequalities.

**BOX:** *What is Already Known on This Topic:* Lower educational attainment is linked to increased risk of adverse pregnancy outcomes, and cardiometabolic risk factors are suspected to mediate some of this effect.

*What This Study Adds:* Our findings from using a two-sample Mendelian randomization approach are in support of a causal relationship between lower educational attainment increasing risk of ectopic pregnancy, hyperemesis gravidarum, gestational diabetes, preeclampsia, preterm birth and offspring low birth weight. A sizeable portion of the effect of educational attainment on ectopic pregnancy, gestational diabetes, preeclampsia and preterm birth is mediated by type 2 diabetes mellitus, body mass index, smoking, high-density lipoprotein cholesterol and systolic blood pressure, while these cardiometabolic risk factors combined explain little of the effect on hyperemesis gravidarum or low birth weight.

*How This Study Might Affect Research, Practice, or Policy:* The effects of socioeconomic inequalities on risk of ectopic pregnancy, gestational diabetes, preeclampsia and preterm birth can be reduced by intervening on type 2 diabetes mellitus, body mass index, smoking, high-density lipoprotein cholesterol and systolic blood pressure.

## INTRODUCTION

Socioeconomic factors – education in particular – are strongly linked to adverse pregnancy outcomes.^1–5^ However, it is challenging to modify an individual’s level of education, and opportunities to seek education are not equally distributed throughout the population. It is therefore of great importance to identify modifiable risk factors through which educational attainment exerts its effects.^6^

Conventional observational studies have identified some potential mediating pathways, and cardiometabolic risk factors that particularly stand out.^1^ For instance, pre-pregnancy body mass index (BMI) and systolic blood pressure (SBP) have been observed to explain most of the association between education and gestational hypertension and preeclampsia.^2,3^ Smoking has been observed to likely mediate some of the potential effect of education on preterm birth^7^ and low birth weight^4^, but not on risk of preeclampsia.^2^ For preterm birth, most of the likely effect of educational attainment remains unexplained.^1^ For yet other pregnancy outcomes, such as ectopic pregnancy, hyperemesis gravidarum, and gestational diabetes mellitus, the role of any mediating pathways downstream of educational attainment remain largely unknown.^5,8,9^ Thus, there is a need for a systematic evaluation of targetable risk factors that may help reduce socioeconomic inequalities in pregnancy outcomes.

There are two other important limitations in the literature on the relationship between educational attainment and pregnancy outcomes. First, many studies lack adjustment for key confounders,^1^ and residual confounding by unmeasured or poorly measured factors may have biased the results.^10^ Second, mediation analyses in traditional observational studies are susceptible to measurement error, such as day-to-day variations of a mediator, which in turn underestimates the mediating effect.^11^

Mendelian randomization (MR) studies use genetic variants as instruments to evaluate the association between an exposure and an outcome. Because genetic variants are allocated at random and not influenced by lifestyle factors and chronic conditions, MR studies are robust to bias by both measured and unmeasured confounders. Furthermore, because genetic variants serve as proxies for the long-term effect of an exposure or mediator, MR studies are generally robust to non-differential measurement error.^12^

We aimed to conduct the first MR study to evaluate the mediating pathways underlying the relationship between educational attainment and pregnancy complications. Specifically, we focused on the following six pregnancy complications and outcomes that are common and/or severe: Ectopic pregnancy, hyperemesis gravidarum, gestational diabetes, preeclampsia, preterm birth, and offspring birth weight. For the mediating factors, our choice of cardiometabolic risk factors was informed by the latest scientific statement by the American Heart Association on optimizing pregnancy health.^13^ In the present study, we therefore investigated the role of type 2 diabetes mellitus (T2DM), BMI, smoking, high-density lipoprotein cholesterol (HDL-C), and SBP in mediating the effect of educational attainment on the risk of pregnancy complications.

## METHODS

### Study Design

In this two-sample MR study we used publicly available, summary-level data with relevant ethical approvals, which did not require institutional review board approval. The paper has been reported according to the STROBE-MR guidelines.^14^

In a two-sample MR study, summary level data from genome-wide association studies (GWASs) are used to find genetic proxies of an exposure, and investigate the associations of these proxies with an outcome.^10^ Specifically, for a given single-nucleotide polymorphism (SNP) used as a genetic proxy – or instrument – the SNP-outcome association is divided by the SNP-exposure association to provide an estimate of the exposure-outcome association, often referred to as the Wald ratio.^10^ Effects across multiple SNPs are pooled using meta-analytical approaches. The steps of a two-sample MR are as follows: First, we identify genetic instruments of the exposure of interest (i.e., educational attainment), next we extract data on how these genetic instruments affect levels of the mediators (e.g., BMI) and outcomes (e.g., preeclampsia), and finally we run analyses as described below.

To limit the risk of confounding due to population stratification, only data from subjects of European ancestry were included.^10^

The summary level data used for this study were publicly available. Data from Steinthorsdottir *et al.*^15^ was obtained through application to the Wellcome Sanger Institute (https://ega-archive.org).

### Instrumental Variable Selection for Educational Attainment

Genetic instruments for education were extracted from a GWAS by Okbay *et al*. (Table 1), the largest GWAS on educational attainment at the time of analysis.^16^ The study evaluated years of education in a combined sample of 3,037,499 subjects. The mean number of years of education was 15.4 years with a standard deviation (SD) of 3.4. We extracted SNPs that were strongly associated with educational attainment, defined as a p-value <5e-8, and that were independent of one-another, defined as a pairwise R^2^ <0.01 based on the 1000 Genomes Project European ancestry superpopulation. The number of SNPs included as genetic instruments for educational attainment varied by outcome under study, from 1,294 for birth weight to 1,727 for T2DM.

**Table 1.**
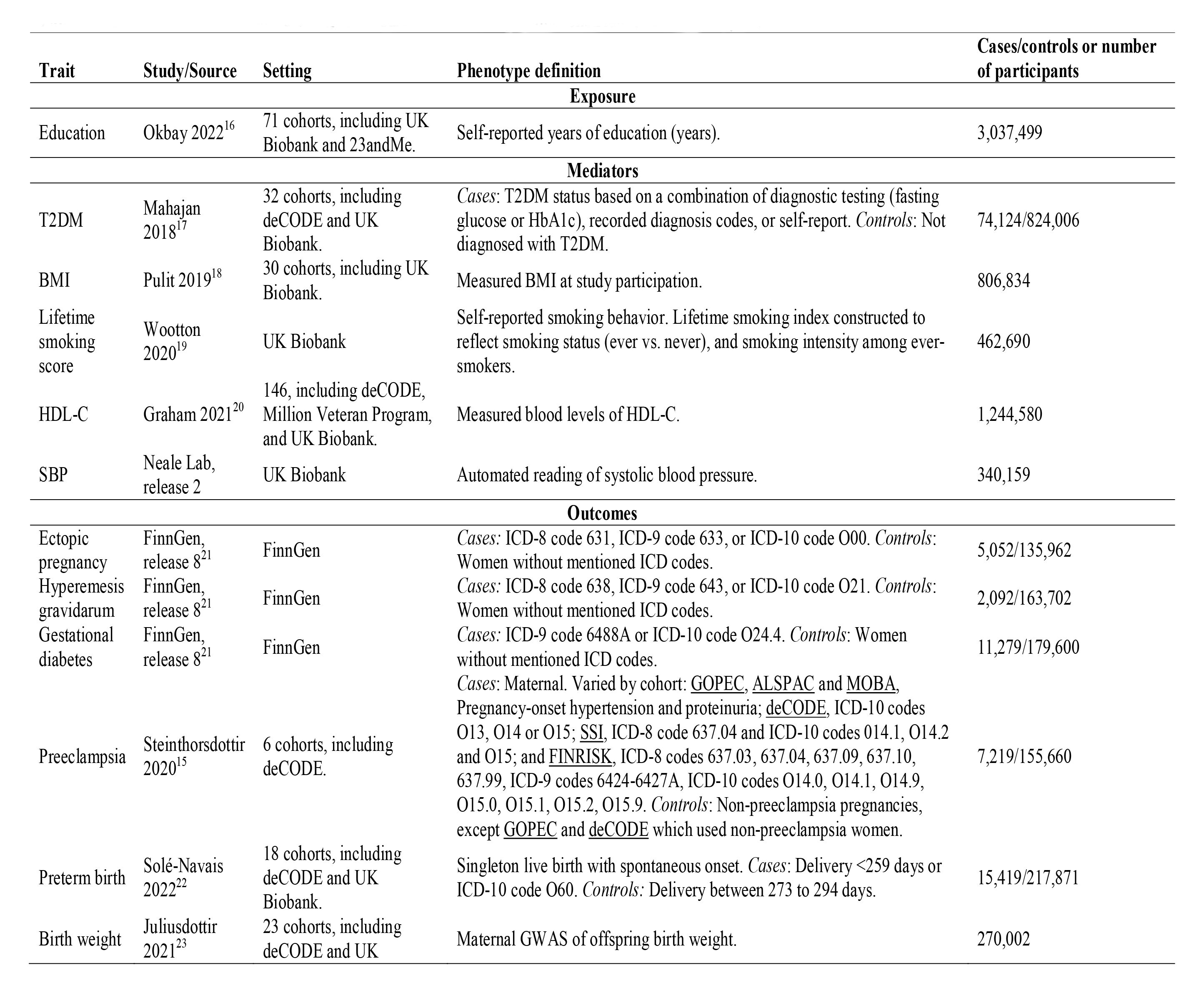
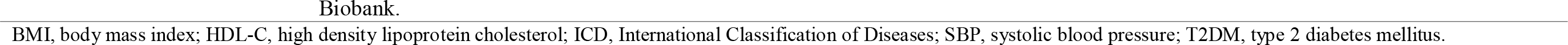
Genome-wide association studies used as sources for two-sample Mendelian randomization analyses.

| Trait | Study/Source | Setting | Phenotype definition | Cases/controls or number of participants |
| --- | --- | --- | --- | --- |
| **Exposure** |  |  |  |  |
| Education | Okbay 2022<sup>16</sup> | 71 cohorts, including UK Biobank and 23andMe. | Self-reported years of education (years). | 3,037,499 |
| **Mediators** |  |  |  |  |
| T2DM | Mahajan 2018<sup>17</sup> | 32 cohorts, including deCODE and UK Biobank. | *Cases:* T2DM status based on a combination of diagnostic testing (fasting glucose or HbA1c), recorded diagnosis codes, or self-report. *Controls:* Not diagnosed with T2DM. | 74,124/824,006 |
| BMI | Pulit 2019<sup>18</sup> | 30 cohorts, including UK Biobank. | Measured BMI at study participation. | 806,834 |
| Lifetime smoking score | Wootton 2020<sup>19</sup> | UK Biobank | Self-reported smoking behavior. Lifetime smoking index constructed to reflect smoking status (ever vs. never), and smoking intensity among ever-smokers. | 462,690 |
| HDL-C | Graham 2021<sup>20</sup> | 146, including deCODE, Million Veteran Program, and UK Biobank. | Measured blood levels of HDL-C. | 1,244,580 |
| SBP | Neale Lab, release 2 | UK Biobank | Automated reading of systolic blood pressure. | 340,159 |
| **Outcomes** |  |  |  |  |
| Ectopic pregnancy | FinnGen, release 8<sup>21</sup> | FinnGen | *Cases:* ICD-8 code 631, ICD-9 code 633, or ICD-10 code O00. *Controls:* Women without mentioned ICD codes. | 5,052/135,962 |
| Hyperemesis gravidarum | FinnGen, release 8<sup>21</sup> | FinnGen | *Cases:* ICD-8 code 638, ICD-9 code 643, or ICD-10 code O21. *Controls:* Women without mentioned ICD codes. | 2,092/163,702 |
| Gestational diabetes | FinnGen, release 8<sup>21</sup> | FinnGen | *Cases:* ICD-9 code 648.8A or ICD-10 code O24.4. *Controls:* Women without mentioned ICD codes. | 11,279/179,600 |
| Preeclampsia | Steinthorsdottir 2020<sup>15</sup> | 6 cohorts, including deCODE. | *Cases:* Maternal. Varied by cohort: <u>GOPEC</u>, <u>ALSPAC</u> and <u>MOBA</u>, Pregnancy-onset hypertension and proteinuria; <u>deCODE</u>, ICD-10 codes O13, O14 or O15; <u>SSI</u>, ICD-8 code 637.04 and ICD-10 codes O14.1, O14.2 and O15; and <u>FINRISK</u>, ICD-8 codes 637.03, 637.04, 637.09, 637.10, 637.99, ICD-9 codes 642.4-642.7A, ICD-10 codes O14.0, O14.1, O14.9, O15.0, O15.1, O15.2, O15.9. *Controls:* Non-preeclampsia pregnancies, except <u>GOPEC</u> and <u>deCODE</u> which used non-preeclampsia women. | 7,219/155,660 |
| Preterm birth | Solé-Navais 2022<sup>22</sup> | 18 cohorts, including deCODE and UK Biobank. | Singleton live birth with spontaneous onset. *Cases:* Delivery <259 days or ICD-10 code O60. *Controls:* Delivery between 273 to 294 days. | 15,419/217,871 |
| Birth weight | Juliusdottir 2021<sup>23</sup> | 23 cohorts, including deCODE and UK | Maternal GWAS of offspring birth weight. | 270,002 |

BMI, body mass index; HDL-C, high density lipoprotein cholesterol; ICD, International Classification of Diseases; SBP, systolic blood pressure; T2DM, type 2 diabetes mellitus.

### Outcomes

Six pregnancy outcomes that are common and/or important contributors to maternal and offspring morbidity and mortality were evaluated. Genetic associations for three of the outcomes – ectopic pregnancy, hyperemesis gravidarum and gestational diabetes – were extracted from a database of publicly available GWAS summary statistics from FinnGen (Table 1).^21^ Results from data freeze 8 were used. For preeclampsia and preterm birth, we used data from studies by Steinthorsdottir *et al.*^15^ and Solé-Navais *et al.*^22^, respectively (Table 1). Finally, for birth weight, we used data from a maternal GWAS of offspring birth weight by Juliusdottir *et al* (Table 1).^23^ All pregnancy outcomes were binary, except from birth weight, which was analyzed as a continuous outcome.

### Mediators

We chose five cardiometabolic traits from the list of targetable risk factors to optimize pregnancy health according to the American Heart Association’s 2023 scientific statement.^13^ Based on our previous research on lipids and risk of preeclampsia,^24^ we decided to evaluate HDL-C instead of non-HDL-C. To limit the number of potential mediators evaluated, and to focus on more specifically modifiable phenotypes that have strong genetic predictors, we did not evaluate sleep health, diet, or physical activity as mediators. SNP-effects on risk of T2DM, BMI and HDL-C were extracted from the studies by Mahajan *et al.*^17^, Pulit *et al.*^18^, and Graham *et al.*^20^, respectively. For smoking behavior, we used a GWAS by Wootton *et al.* on a lifetime smoking index that reflects a combination of smoking status (ever-smoker vs. never-smoker), and smoking duration, heaviness and cessation.^19^ Finally, we used GWAS results from UK Biobank created by the Neale lab (release number 2; http://nealelab.is/uk-biobank/) that evaluated SBP without adjustments for BMI. T2DM was the only binary mediator, while the others were continuous.

### Analyses

In order to address the aims of this study, we evaluated the following measures: 1) The total effect of educational attainment on each pregnancy outcome, 2) the total effect of education on each cardiometabolic risk factor, 3) the total effect of each cardiometabolic risk factor on each pregnancy outcome, and 4) the direct effect of educational attainment on each pregnancy outcome after accounting for each cardiometabolic risk factor separately and combined. The three former analyses were based on univariable MR analyses, while the last analysis was based on multivariable MR. Under the instrumental variable assumptions, the univariable MR estimate represents the total effect of the exposure, whereas the multivariable MR estimate represents the direct effect of the exposure (that is, the effect of intervening on the exposure but holding all mediators constant).

Prior to analyses, we harmonized the files to ensure that the effect estimate of a given SNP was oriented to the same allele in all files.

### Univariable Mendelian Randomization Analyses

For each association, e.g., educational attainment and birth weight, we calculated the Wald ratio per SNP and used inverse-variance weighted (IVW) analysis to summarize the effect of all SNPs, which puts more emphasis on the estimates with the lowest variance.^10^ For the IVW estimate to be unbiased, however, all SNPs included in the analysis must be valid. There are three key assumptions that must be met for an instrument to be valid: It must be associated with the exposure, it cannot be associated with a confounder of the exposure-outcome association, and it is not associated with the outcome other than through the exposure.^10^ Often, a SNP affects multiple biological pathways, and this genetic horizontal pleiotropy can therefore violate the instrumental variable assumptions. For all univariable MR analyses, we therefore conducted three sensitivity analyses that provide unbiased estimates even in the presence of some invalid instruments: The weighted median, weighted mode, and MR Egger regression.^10^ The weighted median and weighted mode approaches calculate a measure of central tendency of the instruments included in the analysis, and respectively assumes that most variants are valid or that more variants estimate the true causal effect than any other quantity.^25^ The MR Egger regression will provide an unbiased estimate if the pleiotropic effects of each variant are independent of the variant-exposure associations.^25^

### Mediation Analyses

We calculated the direct effect of educational attainment on each pregnancy outcome by conducting multivariable MR analyses with each of the five cardiometabolic mediators at a time, and then all mediators combined.^10^ The total effect was provided by the univariable MR analyses as described above. To calculate the proportion mediated (PM), we divided the direct effect by the total effect and subtracted from 1. Finally, we estimated the standard errors using bootstrapping.^26^ The mediation analyses were based on the estimates from the IVW analyses.

### Software

All analyses were run using R (version 4.2.0) and the MendelianRandomization (version 0.7.0), TwoSampleMR (version 0.5.6) and metaphor (version 3.4.0) packages.

### Patient and Public Involvement

We did not involve members of the public or patients when designing the study, interpreting the results, or writing the manuscript; they were not involved in the dissemination plans of this research.

## RESULTS

### Evidence for the Total Effect of Genetically-Predicted Educational Attainment on Pregnancy Outcomes and Cardiometabolic Mediators

The genetic instruments for educational attainment explained 7.5% of its variance, with F-statistics for the individual SNPs ranging from 28 to 576 (median 49). There was a strong protective association of higher level of genetically-predicted educational attainment with all pregnancy outcomes and a strong positive association with offspring birth weight (Figure 1). In terms of risk of the binary pregnancy complications, the protective association of education ranged from odds ratio (OR) 0.53 (95% confidence interval [CI] 0.46 to 0.60) to 0.81 (95% CI 0.71 to 0.93) for ectopic pregnancy and preeclampsia, respectively, per SD increase in genetically-predicted years of education. One SD increase in genetically-predicted years of education was associated with a 41.76 g (95% 27.71 to 55.80) higher birth weight. These associations were robust in sensitivity analyses that evaluated potential bias due to genetic pleiotropy (Supplementary Table 1).

**Figure 1.**
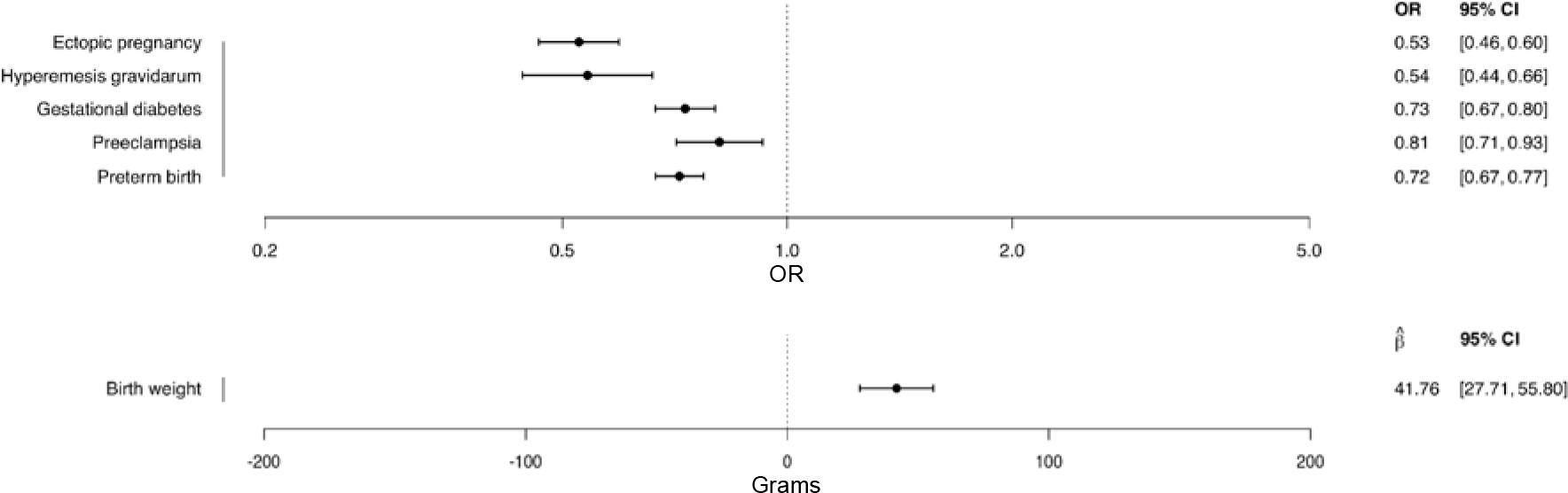
Associations between genetically-predicted educational attainment and pregnancy outcomes. *Legend:* Inverse-variance weighted two-sample Mendelian randomization analyses. Estimates are odds ratios/grams per one standard deviation increase of genetically-predicted years of education (3.4 years). CI, confidence interval; OR, odds ratio.

Educational attainment was also strongly associated with each of the considered cardiometabolic mediators (Figure 2) and this was supported by the sensitivity analyses accounting for pleiotropy (Supplementary Table 2).

**Figure 2.**
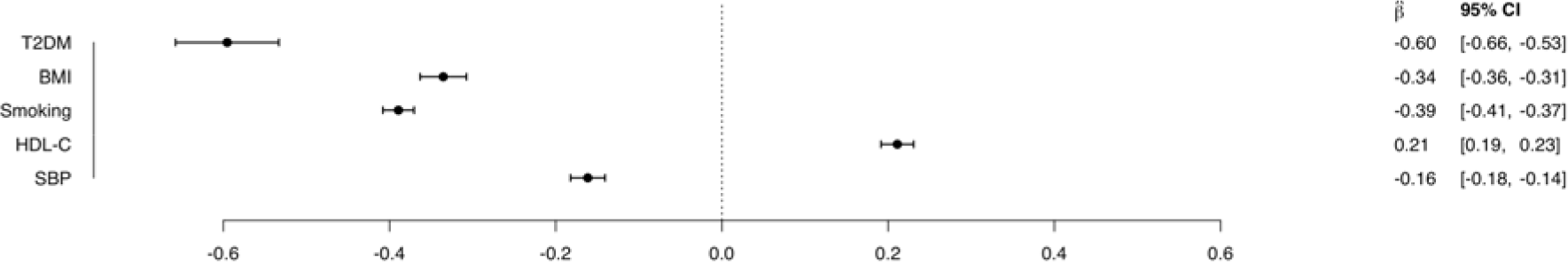
Associations between genetically-predicted educational attainment and cardiometabolic mediators. *Legend:* Inverse-variance weighted two-sample Mendelian randomization analyses. Estimates are the change in the mediator per one standard deviation increase of genetically-predicted years of education (3.4 years), and the mediators are in log(odds) units for T2DM and in standard deviation units for the other cardiometabolic risk factors. BMI, body mass index; CI, confidence interval; HDL-C, high-density lipoprotein cholesterol; OR, odds ratio; SBP, systolic blood pressure; T2DM, type 2 diabetes mellitus.

### The Association between Genetically-Predicted Cardiometabolic Mediators and Pregnancy Outcomes

All genetically-predicted cardiometabolic mediators were associated with at least one pregnancy outcome, but the strength of the associations varied greatly between different outcomes (Figure 3).

**Figure 3.**
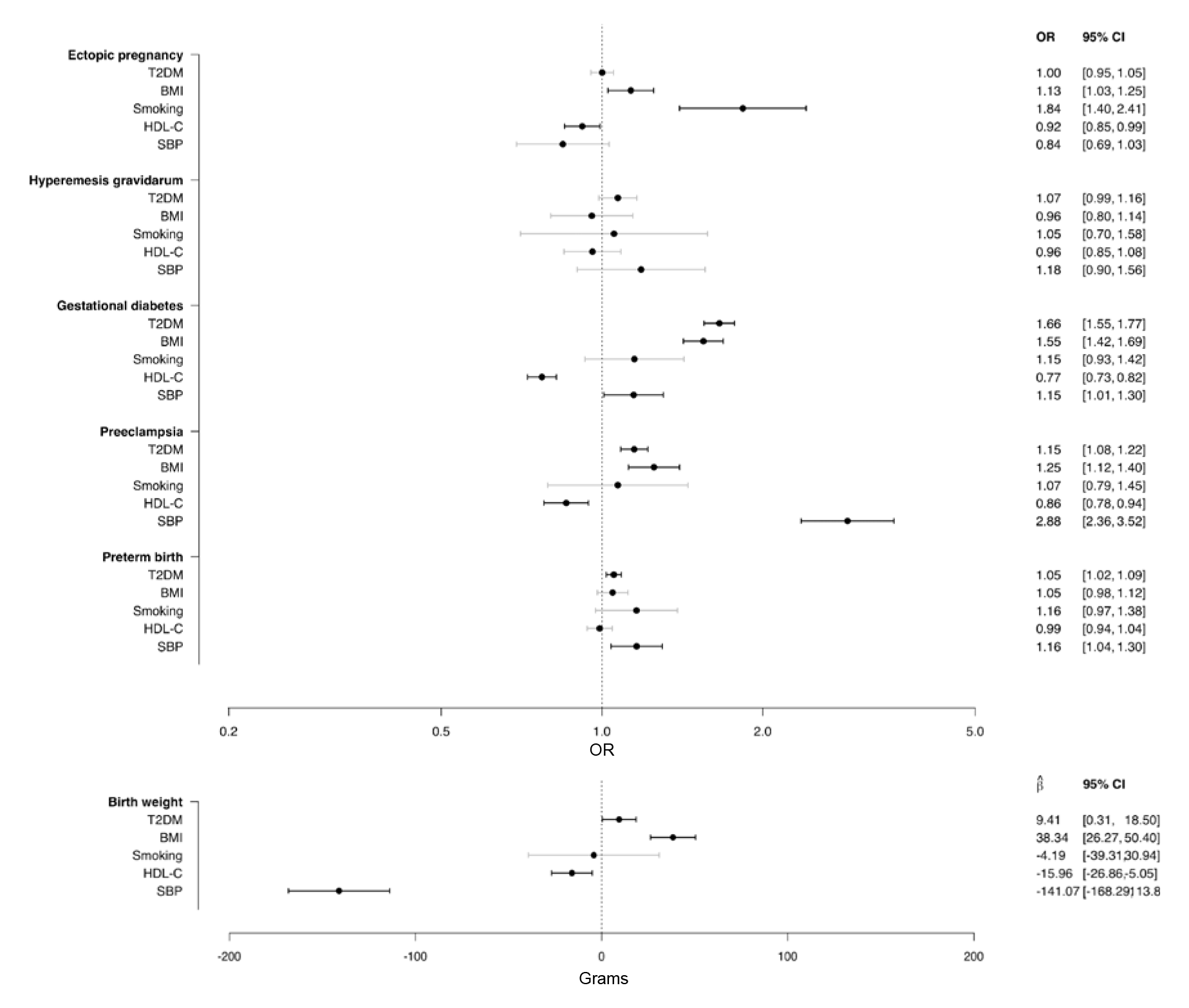
Associations between genetically-predicted cardiometabolic mediators and pregnancy outcomes. *Legend:* Inverse-variance weighted two-sample Mendelian randomization analyses. Estimates are odds ratios/grams per one unit increase of the genetically-predicted log(odds) of T2DM and one standard deviation increase of the other genetically-predicted cardiometabolic traits. BMI, body mass index; CI, confidence interval; HDL-C, high-density lipoprotein cholesterol; OR, odds ratio; SBP, systolic blood pressure; T2DM, type 2 diabetes mellitus.

Genetically-predicted higher T2DM liability was associated with increased risk of gestational diabetes mellitus, preeclampsia, preterm birth, and greater birth weight; genetically-predicted higher BMI was associated with increased risk of ectopic pregnancy, gestational diabetes mellitus, preeclampsia, and greater birth weight; genetically-predicted smoking was associated with an increased risk of ectopic pregnancy; genetically-predicted higher HDL-C was associated with a reduced risk of ectopic pregnancy, gestational diabetes mellitus, preeclampsia, and lower birth weight; and genetically-predicted higher SBP was associated with increased risk of gestational diabetes mellitus, preeclampsia, preterm birth, and lower birth weight. The sensitivity analyses accounting for pleiotropy supported these associations (Supplementary Tables 3 to 7).

### Mediating Pathways Between Genetically-Predicted Level of Education and Pregnancy Outcomes

Figure 4 displays the univariable and multivariable MR estimates of educational attainment on each pregnancy outcome, representing the total effect and direct effect after accounting for each cardiometabolic mediator alone and all combined. We also reported the attenuation in the multivariable MR estimate compared with the univariable MR estimate, labelled as the PM.

**Figure 4.**
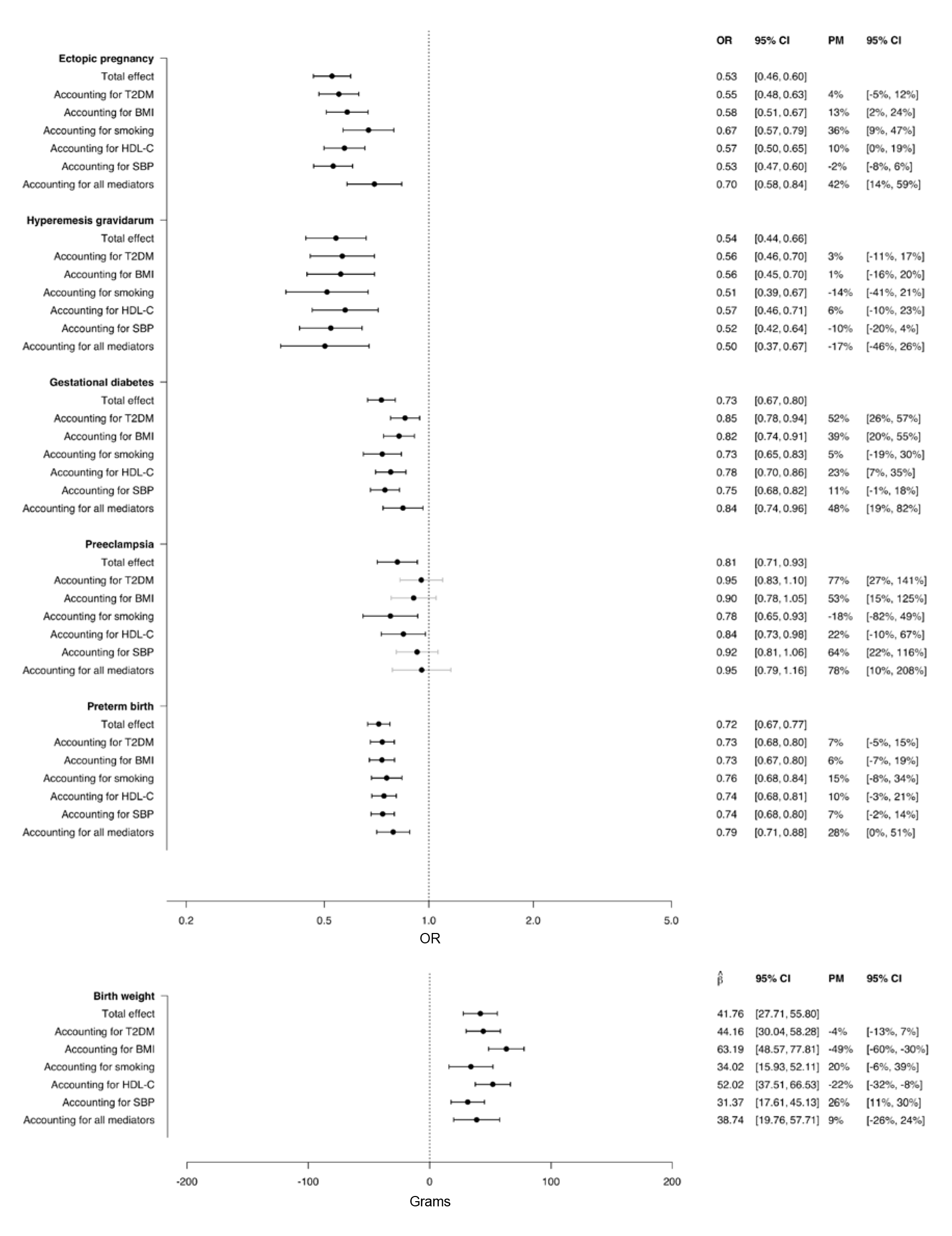
Associations between genetically-predicted educational attainment and pregnancy outcomes after accounting for cardiometabolic mediators. *Legend:* Mendelian randomization mediation analyses. Estimates are odds ratios/grams per one standard deviation increase of genetically-predicted years of education (3.4 years). BMI, body mass index; CI, confidence interval; HDL-C, high-density lipoprotein cholesterol; OR, odds ratio; PM, proportion mediated; SBP, systolic blood pressure; T2DM, type 2 diabetes mellitus.

The attenuation in estimates on adjustment for cardiometabolic traits ranged from almost zero for the association between genetically-predicted education and hyperemesis gravidarum (PM -17% [95% CI -46% to 26%]) to most for the association with risk of preeclampsia (PM 78% [95% CI 10% to 208%]).

The degree to which individual cardiometabolic factors attenuated the association between genetically-predicted educational attainment and the pregnancy outcomes was largely determined by the strength of the association between genetically-predicted education and the mediator (Figure 2), and between the genetically-predicted mediator and the outcome (Figure 3). Thus, for ectopic pregnancy, smoking attenuated 36% of the association (95% CI 9% to 47%); for gestational diabetes mellitus, T2DM liability attenuated 52% of the association (95% CI 26% to 57%); for preeclampsia, T2DM liability, BMI, and SBP attenuated 77% (95% CI 27% to 141%), 53% (95% CI 15% to 125%), and 64% (22% to 116%) of the association, respectively (Figure 4).

For birth weight, the individual cardiometabolic mediators affected associations in competing directions. For instance, because higher genetically-predicted education was associated with lower SBP (Figure 2), and because increasing genetically-predicted SBP was associated with lower birth weight (Figure 3), the association between genetically-predicted education and birth weight was attenuated after accounting for SBP (PM 26% [95% CI 11% to 30%]) (Figure 4). However, since higher genetically-predicted education was associated with lower BMI (Figure 2) and increasing genetically-predicted BMI was associated with higher birth weight (Figure 3), adjustment for BMI strengthened the association between genetically-predicted education and birth weight (PM -49% [95% CI -60% to -30%]) (Figure 4). Thus, there was no net attenuation of the association between genetically-predicted education and birth weight after accounting for all cardiometabolic mediators (PM 9% [95% CI -26% to 24%]).

## DISCUSSION

In this two-sample MR study with mediation analyses, a high genetically-predicted educational attainment was strongly associated with a reduced risk of every pregnancy outcome assessed and with a higher offspring birth weight. For instance, every genetically-predicted 3.4 years longer education halved the risks of ectopic pregnancy and hyperemesis gravidarum. While the cardiometabolic mediators accounted for half and three-quarters of the estimates for gestational diabetes and preeclampsia, respectively, they did not account for any of the estimate for hyperemesis gravidarum. Most of the individual cardiometabolic risk factors affected the association between genetically-predicted education and birth weight, but because their influence was in different directions the combined attenuation was close to null.

### The Effect of Cardiometabolic Traits on Pregnancy Outcomes

In terms of the relationships between cardiometabolic traits and pregnancy outcomes, our study generally supports the findings from previous MR studies but also evaluates several novel associations. In a previous MR study, we observed that genetically-predicted smoking behavior is strongly linked to risk of ectopic pregnancy,^27^ which was reproduced in the current study. For the other adverse pregnancy outcomes, however, there was no clear association with genetically-predicted smoking behavior. While the onset of ectopic pregnancy is a few days after fertilization, the other pregnancy outcomes occur much later in pregnancy. We hypothesize that the null-effect of smoking of the later-onset pregnancy outcomes may in part be because the genetic associations for smoking behavior were derived from a non-pregnant population which may not fully reflect smoking behavior during pregnancy.^28^ For genetically-predicted HDL-C, this is the first MR study to indicate a potential protective effect on risk of ectopic pregnancy, and our study supports previous MR studies that have found evidence of a protective effect on the risk of gestational diabetes and preeclampsia.^24,29^ While a previous MR study observed no association between genetically-predicted HDL-C and offspring birth weight,^30^ we observed a negative association; the discrepancy may be explained by our study using updated genetic datasets evaluating many more subjects. We found a positive association between genetically-predicted BMI and risk of gestational diabetes, preeclampsia, and birth weight, as previously observed.^24,29,31,32^ We also found that high genetically-predicted BMI was associated with an increased risk of ectopic pregnancy; an association that was not significant in a previous MR study by our group due to smaller sample size.^27^ Our study supports previous MR studies reporting a positive association between genetically-predicted SBP and risk of preeclampsia – expected due to shared etiologies^15^ – and lower birth weight,^31^ and provide new data on a positive association with risk of gestational diabetes and risk of preterm birth. Finally, we found that genetically-predicted T2DM liability was associated with an increased risk of gestational diabetes, preeclampsia and higher offspring birth weight as reported in previous MR studies,^31,33^ and a previously unreported association with increased risk of preterm birth.

### Mediating Pathways Between Level of Education and Pregnancy Outcomes

To our knowledge, only one previous MR study has considered the relationship between education and a pregnancy outcome. That study by Liu *et al*. found higher genetically-predicted educational attainment to be associated with increased birth weight, where the strength of association was more pronounced compared with what we found (although our point estimate was within the confidence interval of their analysis).^32^ One reason for the difference may be that we used updated GWASs that were roughly 10 and 3 times larger for educational attainment and birth weight, respectively. While they did not perform formal mediation analyses, they observed comparable results to the main analysis after conducting multivariable MR including BMI and alcohol consumption. This is in contrast to our finding of a stronger association between genetically-predicted educational attainment and birth weight after accounting for BMI. Traditional observational studies have reported that smoking mediates more than a third of the effect of educational attainment on offspring birth weight.^4,34^ This is more pronounced than what our findings suggest, which may be because our smoking variants were imperfect instruments for smoking during pregnancy, as discussed above. Our findings also support previous observations that BMI and smoking mediate in opposite directions, thereby masking socially differentiated healthy fetal growth.^34^

A recent systematic review summarized the literature of conventional observational studies on mediating pathways between socioeconomic status (including educational attainment) and risk of preterm birth.^1^ The individual studies included in the review had discrepant findings but generally reported mediating effects of smoking and BMI comparable to what we found, which is to say fairly small mediated effects. When including T2DM liability, HDL-C and SBP – which were not identified as potential mediators in the review – our analyses support that the five cardiometabolic traits combined may explain roughly a quarter of the effect of genetically-predicted education on risk of preterm birth. The high discrepancy between the individual studies included in the review may partly be due to measurement error of the mediators, which is largely overcome when using genetic instruments as proxies for the mediators.^12^

For preeclampsia and hypertensive disorders of pregnancy, previous studies have observed that the protective effect of high educational attainment disappears after accounting for pre-pregnancy BMI or SBP,^2,3^ which is the same as what we find. Furthermore, smoking behavior has been observed to not mediate any of the effect of educational attainment on risk of gestational hypertension,^2^ which is also supported by our findings. What has not previously been reported is our results suggesting that T2DM liability may mediate the majority of the effect of genetically-predicted education on risk of preeclampsia.

While educational attainment has previously been observed to be strongly linked to risk of gestational diabetes, little has been reported on potential mediating pathways.^5^ Not surprisingly given their shared etiologies,^35^ our findings support that the majority of the effect of genetically-predicted educational attainment may be mediated through T2DM liability.

High genetically-predicted level of education was associated with a reduced risk of hyperemesis gravidarum, similar to findings in a Norwegian register-based study.^8^ Interestingly, although this was one of the most pronounced associations observed in our study, the analyses suggest that it was not mediated by any of the five cardiometabolic traits. In other words, although educational attainment may have a strong effect on risk of hyperemesis gravidarum, this is due to factors other than the prenatal cardiometabolic profile.

Finally, while traditional observational studies have observed that high educational attainment is associated with a reduced risk of ectopic pregnancy,^9^ we are the first to show that almost half of this estimate may be explained by cardiometabolic risk factors, smoking in particular.

### Clinical and Public Health Implications

Our study has several important contributions relevant to policy. First, our study strongly supports previous observations that low levels of education are associated with an increased risk of adverse pregnancy outcomes. This is important, because the triangulation of evidence between traditional observational studies and genetic epidemiological studies – with their different sources of bias – underscores that educational attainment is key for healthy pregnancies at the population level. Policies to facilitate access to continuing and higher education for all may thus improve pregnancy health. However, it can be challenging to improve educational attainment, and among those with low educational attainment, it is crucial to identify targetable factors that may reduce the risk of adverse pregnancy outcomes.

Optimizing all the cardiometabolic traits would considerably lower the risk of ectopic pregnancy, gestational diabetes and preeclampsia, but would have little effect on hyperemesis gravidarum. Subjects with little education and with an otherwise underlying susceptibility to a particular outcome (e.g., previous pelvic surgery leading to an increased risk of ectopic pregnancy) would benefit from more targeted lifestyle changes (e.g., quitting smoking). Finally, for all the evaluated pregnancy outcomes, except preeclampsia, the majority of the effect of educational attainment was mediated through other pathways than the cardiometabolic traits considered. Thus, identifying these other factors will be important in order to reduce the social inequalities in pregnancy health.

### Strengths and Limitations

A major strength of our study is that we applied instrumental variable analyses using genetic instruments which allowed for assessment of the causal role of cardiometabolic mediators in the association between genetically-predicted educational attainment and risk of several pregnancy outcomes. Due to the random allocation of alleles and because these alleles are static throughout a subject’s life, this design is much less likely to be affected by non-differential measurement error of the mediator and confounding compared with traditional observational studies.^10,11^ A potential limitation is that the genetic associations of the mediators were collected from studies that evaluated non-pregnant populations. This may particularly affect behavioral risk factors such as smoking during pregnancy. A study using data from the Avon Longitudinal Study of Parents and Children and the Norwegian Mother, Father and Child Cohort study found that a polygenic risk score for smoking explained 1-3% of variance of smoking during pregnancy.^28^ While this is lower than the 4% explained variance in smoking among non-pregnant subjects observed in the GWAS of which the polygenic risk score was based,^36^ it is still clear that genetic instruments of smoking behavior from a non-pregnant population reflect some of the smoking behavior during pregnancy. Our study is the first MR study to evaluate mediating pathways between educational attainment and pregnancy outcomes. Additionally, it is the first MR study to evaluate most of the associations between educational attainment, the five cardiometabolic traits, and the six pregnancy outcomes. Thereby, using data from large genetic studies and applying robust methods, we were able to present many associations between genetically-predicted educational attainment, cardiometabolic traits and pregnancy outcomes not previously described. To avoid confounding due to population stratification, we evaluated the same genetic ancestry group across all traits.^10^ Consequentially, an important limitation of our study is that we were only able to consider subjects of European ancestry, and we strongly encourage studies evaluating other ancestry groups.

## Conclusion

By using causal genetic epidemiological models, our results suggest that interventions aimed at reducing BMI and SBP, reducing T2DM and smoking prevalence, and increasing HDL-C prior to and during pregnancy would lead to reductions in adverse pregnancy outcomes attributable to lower educational attainment. Except for preeclampsia, most of the effect of genetically-predicted educational attainment on the pregnancy outcomes considered may be mediated through other pathways than these cardiometabolic risk factors, which warrants future studies on additional targetable mediators.

## Data Availability

All data used were publicly available.

