## Supplementary for "Pathways mediating the effect of education on pregnancy outcomes: A Mendelian randomization study"

| **Supplementary Table 1**. Univariable Mendelian randomization analyses of educational attainment on pregnancy outcomes. | | | | | | | | | |
| --- | --- | --- | --- | --- | --- | --- | --- | --- | --- |
|  | **Number  of SNPs** | **Inverse variance weighted** | | **Weighted mode** | | **Weighted median** | | **MR Egger** | |
|  |  | **OR (95% CI)** | **P-value** | **OR (95% CI)** | **P-value** | **OR (95% CI)** | **P-value** | **OR (95% CI)** | **P-value** |
| Ectopic pregnancy | 1640 | 0.53 (0.46, 0.60) | 2.78E-24 | 0.47 (0.23, 0.94) | 3.29E-02 | 0.51 (0.42, 0.61) | 1.98E-12 | 0.42 (0.28, 0.62) | 1.46E-05 |
| Hyperemesis gravidarum | 1640 | 0.54 (0.44, 0.66) | 1.69E-09 | 1.13 (0.31, 4.05) | 8.56E-01 | 0.64 (0.47, 0.86) | 2.96E-03 | 0.65 (0.34, 1.23) | 1.88E-01 |
| Gestational diabetes | 1639 | 0.73 (0.67, 0.80) | 2.23E-11 | 0.73 (0.44, 1.22) | 2.28E-01 | 0.74 (0.65, 0.84) | 5.44E-06 | 0.73 (0.54, 0.98) | 3.58E-02 |
| Preeclampsia | 1707 | 0.81 (0.71, 0.93) | 2.08E-03 | 0.76 (0.33, 1.72) | 5.04E-01 | 0.83 (0.68, 1.00) | 5.37E-02 | 0.93 (0.61, 1.42) | 7.26E-01 |
| Preterm birth | 1689 | 0.72 (0.67, 0.77) | 1.24E-18 | 1.09 (0.65, 1.81) | 7.53E-01 | 0.78 (0.70, 0.87) | 1.68E-05 | 0.63 (0.50, 0.80) | 1.36E-04 |
|  |  | **Grams (95% CI)** | **P-value** | **Grams (95% CI)** | **P-value** | **Grams (95% CI)** | **P-value** | **Grams (95% CI)** | **P-value** |
| Birth weight | 1294 | 41.76 (27.71, 55.80) | 5.59E-09 | 125.39 (60.66, 190.12) | 1.53E-04 | 55.85 (39.56, 72.15) | 1.83E-11 | 18.57 (-25.42, 62.56) | 4.08E-01 |
| CI, confidence interval; OR, odds ratio; SNPs, single-nucleotide polymorphisms. | | | | | | | | | |

| **Supplementary Table 2**. Univariable Mendelian randomization analyses of educational attainment on cardiometabolic mediators. | | | | | | | | | |
| --- | --- | --- | --- | --- | --- | --- | --- | --- | --- |
|  | **Number  of SNPs** | **Inverse variance weighted** | | **Weighted mode** | | **Weighted median** | | **MR Egger** | |
|  |  | **Beta (95% CI)** | **P-value** | **Beta (95% CI)** | **P-value** | **Beta (95% CI)** | **P-value** | **Beta (95% CI)** | **P-value** |
| T2DM | 1727 | -0.60 (-0.66, -0.53) | 1.02E-78 | -0.49 (-0.80, -0.19) | 1.66E-03 | -0.58 (-0.64, -0.51) | 2.00E-72 | -0.73 (-0.92, -0.53) | 9.78E-13 |
| BMI | 1724 | -0.34 (-0.36, -0.31) | 5.53E-123 | -0.31 (-0.43, -0.20) | 1.88E-07 | -0.29 (-0.32, -0.27) | 1.86E-166 | -0.39 (-0.48, -0.30) | 3.05E-17 |
| Smoking | 1725 | -0.39 (-0.41, -0.37) | 0 | -0.35 (-0.45, -0.25) | 4.21E-12 | -0.36 (-0.38, -0.34) | 6.19E-254 | -0.36 (-0.42, -0.31) | 2.53E-32 |
| HDL-C | 1725 | 0.21 (0.19, 0.23) | 3.93E-99 | 0.22 (0.14, 0.29) | 1.78E-08 | 0.20 (0.19, 0.22) | 3.21E-143 | 0.29 (0.23, 0.35) | 1.73E-19 |
| SBP | 1724 | -3.12 (-3.53, -2.72) | 1.70E-52 | -3.73 (-5.63, -1.83) | 1.21E-04 | -2.96 (-3.40, -2.52) | 7.90E-40 | -1.97 (-3.24, -0.70) | 2.37E-03 |
| Betas reflect standard deviation units for all traits except for T2DM where it reflects log(odds ratio). BMI, body mass index; CI, confidence interval; HDL-C, high-density lipoprotein cholesterol; SBP, systolic blood pressure; SNPs, single-nucleotide polymorphisms; T2DM, type 2 diabetes mellitus. | | | | | | | | | |

| **Supplementary Table 3**. Univariable Mendelian randomization analyses of type 2 diabetes mellitus on pregnancy outcomes. | | | | | | | | | |
| --- | --- | --- | --- | --- | --- | --- | --- | --- | --- |
|  | **Number  of SNPs** | **Inverse variance weighted** | | **Weighted mode** | | **Weighted median** | | **MR Egger** | |
|  |  | **OR (95% CI)** | **P-value** | **OR (95% CI)** | **P-value** | **OR (95% CI)** | **P-value** | **OR (95% CI)** | **P-value** |
| Ectopic pregnancy | 237 | 1.00 (0.95, 1.05) | 9.74E-01 | 0.95 (0.83, 1.08) | 4.52E-01 | 0.98 (0.91, 1.06) | 6.16E-01 | 0.97 (0.86, 1.08) | 5.40E-01 |
| Hyperemesis gravidarum | 237 | 1.07 (0.99, 1.16) | 1.03E-01 | 1.07 (0.87, 1.31) | 5.41E-01 | 1.03 (0.90, 1.18) | 6.46E-01 | 1.15 (0.95, 1.39) | 1.51E-01 |
| Gestational diabetes | 237 | 1.66 (1.55, 1.77) | 1.30E-51 | 1.59 (1.46, 1.74) | 8.19E-21 | 1.60 (1.50, 1.71) | 4.70E-45 | 1.89 (1.63, 2.19) | 6.46E-15 |
| Preeclampsia | 249 | 1.15 (1.08, 1.22) | 3.37E-06 | 1.13 (0.99, 1.29) | 7.75E-02 | 1.14 (1.03, 1.25) | 1.00E-02 | 1.17 (1.02, 1.34) | 3.02E-02 |
| Preterm birth | 244 | 1.05 (1.02, 1.09) | 2.14E-03 | 0.99 (0.91, 1.08) | 8.03E-01 | 1.03 (0.98, 1.09) | 2.61E-01 | 1.08 (1.00, 1.17) | 3.98E-02 |
|  |  | **Grams (95% CI)** | **P-value** | **Grams (95% CI)** | **P-value** | **Grams (95% CI)** | **P-value** | **Grams (95% CI)** | **P-value** |
| Birth weight | 185 | 9.41 (0.31, 18.50) | 4.26E-02 | 17.04 (1.45, 32.63) | 3.34E-02 | 10.88 (2.92, 18.84) | 7.40E-03 | 13.66 (-6.58, 33.89) | 1.88E-01 |
| CI, confidence interval; OR, odds ratio; SNPs, single-nucleotide polymorphisms. | | | | | | | | | |

| **Supplementary Table 4**. Univariable Mendelian randomization analyses of body mass index on pregnancy outcomes. | | | | | | | | | |
| --- | --- | --- | --- | --- | --- | --- | --- | --- | --- |
|  | **Number  of SNPs** | **Inverse variance weighted** | | **Weighted mode** | | **Weighted median** | | **MR Egger** | |
|  |  | **OR (95% CI)** | **P-value** | **OR (95% CI)** | **P-value** | **OR (95% CI)** | **P-value** | **OR (95% CI)** | **P-value** |
| Ectopic pregnancy | 1035 | 1.13 (1.03, 1.25) | 1.31E-02 | 0.83 (0.49, 1.40) | 4.86E-01 | 1.03 (0.89, 1.20) | 6.99E-01 | 1.05 (0.78, 1.40) | 7.68E-01 |
| Hyperemesis gravidarum | 1035 | 0.96 (0.80, 1.14) | 6.27E-01 | 0.98 (0.47, 2.01) | 9.46E-01 | 0.95 (0.74, 1.21) | 6.63E-01 | 0.64 (0.38, 1.09) | 1.05E-01 |
| Gestational diabetes | 1035 | 1.55 (1.42, 1.69) | 1.59E-23 | 1.75 (1.17, 2.61) | 6.21E-03 | 1.59 (1.43, 1.78) | 1.26E-16 | 1.59 (1.23, 2.05) | 4.27E-04 |
| Preeclampsia | 1055 | 1.25 (1.12, 1.40) | 6.94E-05 | 1.17 (0.65, 2.11) | 5.93E-01 | 1.17 (1.00, 1.38) | 5.16E-02 | 1.15 (0.83, 1.59) | 4.12E-01 |
| Preterm birth | 1046 | 1.05 (0.98, 1.12) | 1.72E-01 | 0.86 (0.63, 1.17) | 3.39E-01 | 0.99 (0.89, 1.09) | 7.70E-01 | 0.80 (0.66, 0.97) | 2.38E-02 |
|  |  | **Grams (95% CI)** | **P-value** | **Grams (95% CI)** | **P-value** | **Grams (95% CI)** | **P-value** | **Grams (95% CI)** | **P-value** |
| Birth weight | 834 | 38.34 (26.27, 50.40) | 4.73E-10 | 75.21 (37.02, 113.40) | 1.22E-04 | 43.40 (28.99, 57.82) | 3.61E-09 | 26.35 (-7.60, 60.31) | 1.29E-01 |
| CI, confidence interval; OR, odds ratio; SNPs, single-nucleotide polymorphisms. | | | | | | | | | |

| **Supplementary Table 5**. Univariable Mendelian randomization analyses of smoking on pregnancy outcomes. | | | | | | | | | |
| --- | --- | --- | --- | --- | --- | --- | --- | --- | --- |
|  | **Number  of SNPs** | **Inverse variance weighted** | | **Weighted mode** | | **Weighted median** | | **MR Egger** | |
|  |  | **OR (95% CI)** | **P-value** | **OR (95% CI)** | **P-value** | **OR (95% CI)** | **P-value** | **OR (95% CI)** | **P-value** |
| Ectopic pregnancy | 139 | 1.84 (1.40, 2.41) | 1.26E-05 | 1.62 (0.66, 4.00) | 2.95E-01 | 1.71 (1.15, 2.54) | 7.58E-03 | 1.84 (0.59, 5.79) | 2.97E-01 |
| Hyperemesis gravidarum | 139 | 1.05 (0.70, 1.58) | 8.01E-01 | 0.43 (0.09, 2.06) | 2.93E-01 | 0.96 (0.52, 1.74) | 8.84E-01 | 0.72 (0.13, 3.94) | 7.10E-01 |
| Gestational diabetes | 139 | 1.15 (0.93, 1.42) | 1.99E-01 | 1.13 (0.60, 2.11) | 7.12E-01 | 1.13 (0.86, 1.49) | 3.64E-01 | 0.80 (0.33, 1.95) | 6.19E-01 |
| Preeclampsia | 144 | 1.07 (0.79, 1.45) | 6.58E-01 | 1.67 (0.57, 4.93) | 3.55E-01 | 1.16 (0.75, 1.79) | 5.01E-01 | 2.06 (0.62, 6.85) | 2.42E-01 |
| Preterm birth | 142 | 1.16 (0.97, 1.38) | 9.88E-02 | 1.07 (0.59, 1.94) | 8.36E-01 | 1.13 (0.88, 1.44) | 3.32E-01 | 1.40 (0.68, 2.88) | 3.58E-01 |
|  |  | **Grams (95% CI)** | **P-value** | **Grams (95% CI)** | **P-value** | **Grams (95% CI)** | **P-value** | **Grams (95% CI)** | **P-value** |
| Birth weight | 116 | -4.19 (-39.31, 30.94) | 8.15E-01 | -73.07 (-143.15, -2.99) | 4.33E-02 | -38.72 (-74.21, -3.22) | 3.25E-02 | -63.65 (-198.01, 70.71) | 3.55E-01 |
| CI, confidence interval; OR, odds ratio; SNPs, single-nucleotide polymorphisms. | | | | | | | | | |

| **Supplementary Table 6**. Univariable Mendelian randomization analyses of high-density lipoprotein cholesterol on pregnancy outcomes. | | | | | | | | | |
| --- | --- | --- | --- | --- | --- | --- | --- | --- | --- |
|  | **Number  of SNPs** | **Inverse variance weighted** | | **Weighted mode** | | **Weighted median** | | **MR Egger** | |
|  |  | **OR (95% CI)** | **P-value** | **OR (95% CI)** | **P-value** | **OR (95% CI)** | **P-value** | **OR (95% CI)** | **P-value** |
| Ectopic pregnancy | 1031 | 0.92 (0.85, 0.99) | 2.98E-02 | 1.05 (0.91, 1.21) | 5.23E-01 | 1.04 (0.92, 1.19) | 5.19E-01 | 0.99 (0.88, 1.11) | 8.51E-01 |
| Hyperemesis gravidarum | 1031 | 0.96 (0.85, 1.08) | 5.10E-01 | 1.07 (0.86, 1.33) | 5.64E-01 | 1.08 (0.87, 1.36) | 4.83E-01 | 1.13 (0.94, 1.36) | 1.97E-01 |
| Gestational diabetes | 1031 | 0.77 (0.73, 0.82) | 4.59E-16 | 0.98 (0.88, 1.09) | 6.87E-01 | 0.91 (0.82, 1.00) | 4.57E-02 | 0.96 (0.88, 1.06) | 4.15E-01 |
| Preeclampsia | 1077 | 0.86 (0.78, 0.94) | 1.63E-03 | 0.84 (0.71, 0.98) | 3.09E-02 | 0.81 (0.70, 0.94) | 5.90E-03 | 0.94 (0.81, 1.09) | 4.07E-01 |
| Preterm birth | 1066 | 0.99 (0.94, 1.04) | 7.14E-01 | 1.01 (0.92, 1.11) | 7.97E-01 | 1.01 (0.92, 1.11) | 8.69E-01 | 1.00 (0.92, 1.09) | 9.69E-01 |
|  |  | **Grams (95% CI)** | **P-value** | **Grams (95% CI)** | **P-value** | **Grams (95% CI)** | **P-value** | **Grams (95% CI)** | **P-value** |
| Birth weight | 847 | -15.96 (-26.86, -5.05) | 4.14E-03 | -6.60 (-20.53, 7.33) | 3.53E-01 | -10.77 (-23.51, 1.97) | 9.76E-02 | -3.84 (-20.36, 12.67) | 6.48E-01 |
| CI, confidence interval; OR, odds ratio; SNPs, single-nucleotide polymorphisms. | | | | | | | | | |

| **Supplementary Table 7**. Univariable Mendelian randomization analyses of systolic blood pressure on pregnancy outcomes. | | | | | | | | | |
| --- | --- | --- | --- | --- | --- | --- | --- | --- | --- |
|  | **Number  of SNPs** | **Inverse variance weighted** | | **Weighted mode** | | **Weighted median** | | **MR Egger** | |
|  |  | **OR (95% CI)** | **P-value** | **OR (95% CI)** | **P-value** | **OR (95% CI)** | **P-value** | **OR (95% CI)** | **P-value** |
| Ectopic pregnancy | 253 | 0.84 (0.69, 1.03) | 9.76E-02 | 0.70 (0.34, 1.46) | 3.47E-01 | 0.91 (0.70, 1.19) | 4.87E-01 | 0.87 (0.45, 1.66) | 6.65E-01 |
| Hyperemesis gravidarum | 253 | 1.18 (0.90, 1.56) | 2.31E-01 | 1.10 (0.41, 2.93) | 8.55E-01 | 1.17 (0.79, 1.74) | 4.39E-01 | 1.40 (0.57, 3.44) | 4.62E-01 |
| Gestational diabetes | 253 | 1.15 (1.01, 1.30) | 3.68E-02 | 1.20 (0.81, 1.80) | 3.66E-01 | 1.19 (1.00, 1.42) | 5.21E-02 | 0.97 (0.64, 1.47) | 8.73E-01 |
| Preeclampsia | 265 | 2.88 (2.36, 3.52) | 2.85E-25 | 1.96 (0.86, 4.48) | 1.11E-01 | 2.66 (2.05, 3.45) | 1.51E-13 | 4.95 (2.56, 9.59) | 3.49E-06 |
| Preterm birth | 266 | 1.16 (1.04, 1.30) | 8.03E-03 | 0.88 (0.60, 1.29) | 5.12E-01 | 1.13 (0.97, 1.32) | 1.26E-01 | 0.97 (0.69, 1.39) | 8.86E-01 |
|  |  | **Grams (95% CI)** | **P-value** | **Grams (95% CI)** | **P-value** | **Grams (95% CI)** | **P-value** | **Grams (95% CI)** | **P-value** |
| Birth weight | 230 | -141.07 (-168.29, -113.84) | 3.12E-24 | -127.56 (-172.21, -82.91) | 6.12E-08 | -124.99 (-147.27, -102.71) | 3.98E-28 | -176.96 (-264.44, -89.48) | 9.83E-05 |
| CI, confidence interval; OR, odds ratio; SNPs, single-nucleotide polymorphisms. | | | | | | | | | |
